# Use of Registry Data to Improve Center Outcomes and Inform Global Care Standards in Adult Cystic Fibrosis

**DOI:** 10.64898/2026.04.28.26351666

**Authors:** Rajeshree Joshi, Samantha Lazaro, Sharad Purohit, Kathleen McKie, Caralee Forseen, Varsha Taskar

## Abstract

**Aim:** Cystic fibrosis (CF) care has been transformed by CFTR modulator therapies, yet most efficacy data arise from clinical trials with restrictive eligibility criteria. Real-world registry data can capture treatment outcomes in broader, more diverse patient populations. We used the Cystic Fibrosis Foundation Patient Registry (CFFPR) data to evaluate longitudinal clinical outcomes, and care benchmarking at a single Adult CF Program Center over a decade.

**Methods:** A retrospective, descriptive analysis of CFFPR data (2011–2022) was performed to assess trends in modulator use, lung function (ppFEV1), body mass index (BMI), respiratory microbiology, and pulmonary exacerbations (PEx). Comparative Effectiveness Research (CER) methods were applied to compare outcomes across peak modulator eras: pre-modulator (2011), ivacaftor (2015), mixed-modulator (2019), and elexacaftor/tezacaftor/ivacaftor (ELE/TEZ/IVA) (2021). Program-level outcomes were benchmarked against national network metrics to assess adherence to guideline-based care.

**Results:** Over ten years, median ppFEV_1_ improved from 63.4% (2011) to 78.8% (2021), and BMI increased from 22.3 to 24.8 kg/m^2^. The proportion of adults experiencing more than one PEx annually declined from 39.7% to 19.5%, while *Pseudomonas aeruginosa* (P.a.) culture positivity decreased from 79% to 47%. ELE/TEZ/IVA therapy was associated with greatest clinical improvements. Program-level performance remained comparable to national network benchmarks, reflecting high adherence to standard care metrics.

**Conclusion:** Registry-based CER provides valuable real-world insights into CF care effectiveness and quality improvement. This decade-long analysis demonstrates significant clinical gains associated with modulator therapies and highlights the importance of patient registries in monitoring outcomes, benchmarking care, and informing global CF care models and standards for rare disease management.

**Key Messages:**

□ Real-world registry data enables decade-long evaluation of CFTR modulator effectiveness.
□ Elexacaftor/tezacaftor/ivacaftor demonstrates the greatest clinical benefit among modulator therapies.
□ Benchmarking Adult CF Program performance against the Network of Adult CF Programs facilitates quality improvement and standard care guideline adherence.
□ Patient registries provide insights for personalized care, program-level decision-making, and international standards for rare disease management.

## INTRODUCTION

Cystic fibrosis (CF) is a rare autosomal recessive genetic disorder caused by mutations in the *cystic fibrosis transmembrane conductance regulator* (*CFTR*) gene. This translates into a defective protein involved in chloride transport, thickened secretions, and progressive multisystem involvement. The mutation (F508del) primarily affects the lungs, pancreas, and gastrointestinal tract, leading to chronic respiratory infections, malnutrition, and complications such as CF-related diabetes and osteoporosis.[1-3]

Over the past decade, CF care has been revolutionized by the advent of CFTR modulator therapies that target the underlying protein dysfunction. The first CFTR potentiator, ivacaftor, received approval in 2012, followed by combination therapies—lumacaftor/ivacaftor (LUM/IVA) in 2015, tezacaftor/ivacaftor (TEZ/IVA) in 2018, and the highly effective triple combination ELE/TEZ/IVA in 2019. These modulators have markedly improved lung function, nutritional status, and quality of life for people with CF.[4]

Most evidence supporting modulator efficacy and safety is derived from randomized clinical trials with strict inclusion and exclusion criteria, limiting generalizability to the broader CF population.[5, 6] Real-world data from patient registries can overcome these limitations by capturing outcomes across diverse clinical settings and patient subgroups.[7, 8] The CFFPR in the United States is a robust, longitudinal database that collects standardized information from CF care centers nationwide [9]. These data enable CER to evaluate treatment impact, care variability, and quality improvement opportunities in real-world practice. The present study uses CFFPR data to assess a decade of real-world trends (2011–2022) in CFTR modulator therapy use, clinical outcomes, and program-level adherence to care guidelines in an adult CF population [9]. By combining CER methodology with benchmarking against the national Adult CF Program Network, this study provides longitudinal insights into modulator effectiveness and highlights the potential of registry-based research to inform evidence-driven CF care globally.

## METHODS

### CF Registry Data Abstraction

CF individuals taking standard care treatment at Adult CF Program have consented to the CFF Patient Registry. Each visit data is entered into CFF’s PortCF web portal and stored in the central database CFFPR. The program-level data are compiled annually through the Annual Program Specific Reports, which also provide aggregated network-level metrics for benchmarking. Patient demographic data are collected from each CF Program’s PortCF web portal and include age, sex, race, and CF genotype. The Program-level and the Network-level data are collected for 2011-22 from the registry’s Annual reports for 2020, 2021, and 2022.[10-12] The clinical outcome data extracted includes pulmonary therapy usages, CF-related complications, modulator therapy usage, lung function (ppFEV1), nutritional status via BMI, PEx episodes, and microbiology results from the respiratory cultures. The standard clinical care practices data include annual review metrics for patients evaluated by respiratory therapist, physical therapist, mental health coordinator, dietitian and patients with 1 visit, 1 culture, and 1 pulmonary function test (PFT) done.[10-12] These data provided the basis for longitudinal analysis of modulator trends, clinical outcomes, and program-level care delivery relative to the network.

The study was approved under the data use agreement of the CFFPR and conducted in accordance with the Declaration of Helsinki (1997). All patients provided written informed consent for inclusion in the registry. Institutional Review Board (IRB) approval was not required for analysis of de-identified, aggregated registry data.

### Statistical Analysis

Categorical data is presented as counts and percentages. Differences between the groups for categorical variables were assessed using chi-square tests. Continuous data is presented as means and standard deviations; differences between groups were tested using univariate t-tests. Due to the low number of patients, corrections for multiple testing were not performed.

The clinical and demographic data was stored and processed using Microsoft Excel© (ver MS office 365). Chi-square tests and t-tests were performed in Microsoft Excel. All analyses were focused on visualizing trends over time; no causal inference testing was performed due to the observational nature of registry data.

## RESULTS

### Program-Level Demographics and Genotype Distribution

In 2022, the Adult CF Program population included 102 patients: 82% non-Hispanic White, 14% Black or African-American, and 2% each Asian and Hispanic. The cohort was evenly distributed by sex (50% male, 50% female), with age ranges of 20–75 years for males and 20–82 years for females.

Genotype analysis revealed 40% were F508del-homozygous, 44% F508del-heterozygous, 14% with other known mutations, and 2% with unknown mutations. According to CFTR2 classification, 91% of patients were eligible for one or more modulators: 13 for IVA, 41 for LUM/IVA, 10 for TEZ/IVA, and 93 for ELE/TEZ/IVA based on specific responsive mutations.

### Clinical Characteristics and Comorbidities

Registry reports from 2022 identified pancreatic insufficiency as the most prevalent comorbidity (88.5%). Other complications included cystic fibrosis–related diabetes (42.5%), depression (28.7%), anxiety disorder (26.4%), advanced lung disease (5.7%), osteoporosis (13.8%), allergic bronchopulmonary aspergillosis (6.9%), distal intestinal obstruction syndrome (2.3%), liver cirrhosis (2.3%), and five patients had undergone bilateral lung transplantation.

### Pulmonary Therapies and Standard Care Practices

The use of airway clearance and inhaled therapies was consistent with national standards. In 2022, Dornase Alpha was prescribed in 88.5%, hypertonic saline in 78.2%, inhaled steroids in 40.9%, and exercise-based airway clearance in 5.7%. The most common anti-pseudomonal regimens included azithromycin (79.4%), tobramycin (55.6%), and inhaled aztreonam (29.7%).

Adherence to annual care metrics remained high, with more than 93% of adults completing at least one annual visit, respiratory culture, and pulmonary function test (PFT), also meeting the standard annual evaluations by the multi-disciplinary care team including the respiratory therapist, mental health coordinator, nutritionist, social worker and pharmacist. Standard care, screening and prevention procedures (including OGTT screenings in non-diabetics, DEXA scans, liver enzyme measured, fat-soluble vitamins measured, influenza vaccination, tobacco smoke exposure evaluation) showed increasing compliance over the study period, aligning closely with network benchmarks.

### Lung Function and Nutritional Outcomes

Longitudinal analysis demonstrated steady improvement in lung function (ppFEV_1_) and nutritional status over the decade (Figure 1). Median ppFEV_1_ increased from 63.4% in 2011 to 78.8% in 2021, while median BMI rose from 22.3 kg/m^2^ to 24.8 kg/m^2^. At each modulator peak usage, ppFEV_1_ improved from 63.4% (2011, non-modulator baseline) to 68.3% (2015, IVA era), 70.5% (2019, mixed modulator cohort), and 78.8% (2021, ELE/TEZ/IVA). BMI increased in parallel from 22.3 → 23.0 → 23.6 → 24.8 kg/m^2^ across the same periods. These trends reflect improved outcomes with CFTR modulator therapy, with ELE/TEZ/IVA showing higher effectiveness than IVA. Table 2 highlights the progression across the selected modulator peak usage time points.

**Figure 1.**
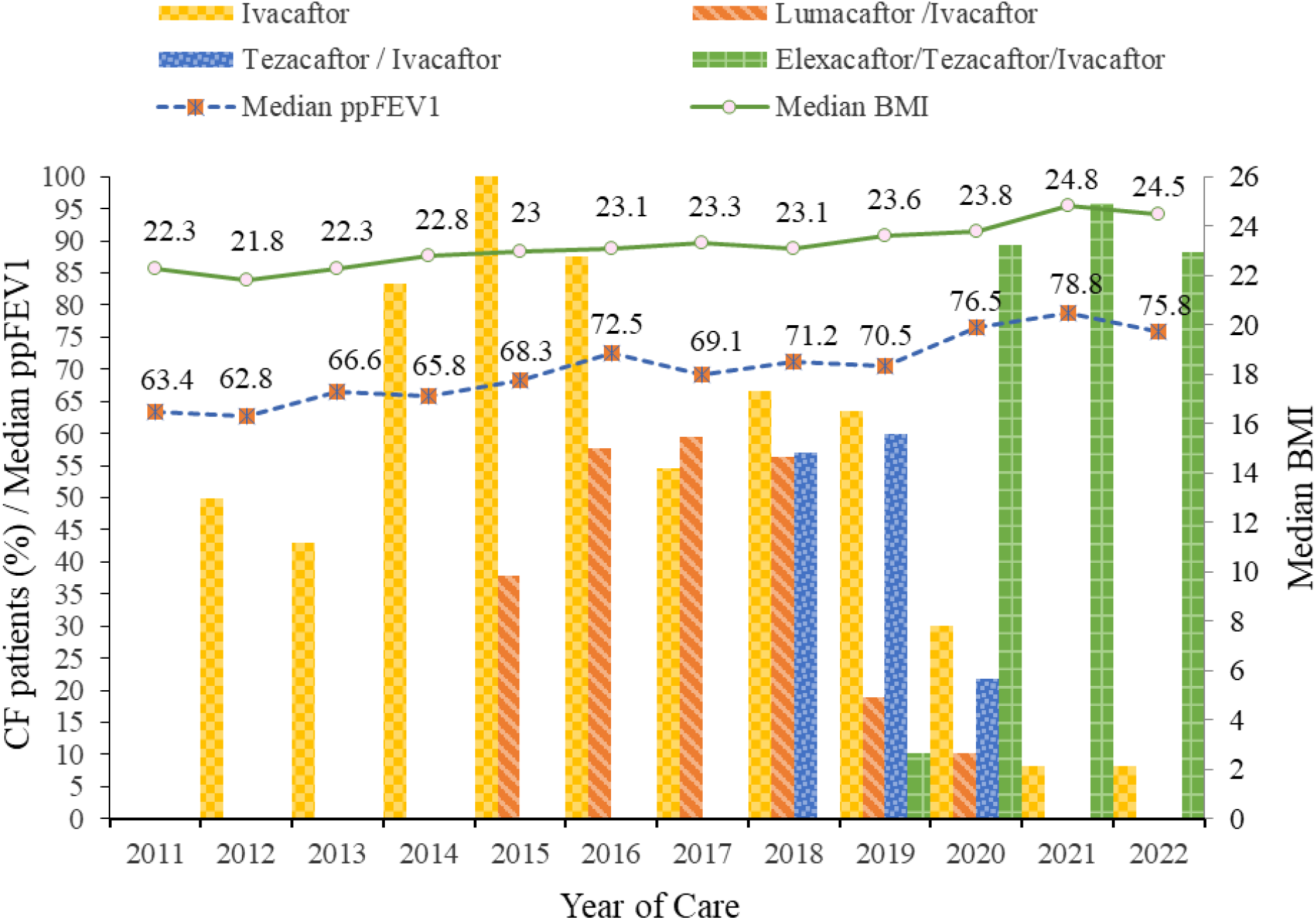
Comparative effectiveness of CFTR modulators on clinical outcomes in adults at our Adult CF Program, 2011-2022. Vertical bars represent the years of ivacaftor (IVA), lumacaftor/ivacaftor (LUM/IVA), tezacaftor/ivacaftor (TEZ/IVA), and elexacaftor/tezacaftor/ivacaftor (ELE/TEZ/IVA) availability. Longitudinal trends in lung function (percent predicted forced expiratory volume in 1 second, ppFEV_1_) and body mass index (BMI) are displayed as lines with markers.

**Table 1.** Comparative effectiveness analysis of CFTR modulators on clinical outcomes in adult cystic fibrosis patients at our program, based on Cystic Fibrosis Foundation Patient Registry data from 2011-2022. Clinical outcomes during peak use of elexacaftor/tezacaftor/ivacaftor (ELE/TEZ/IVA) in 2021 are compared with ivacaftor (IVA) in 2015, a mixed-modulator cohort in 2019, and the pre-modulator baseline year 2011.

| Variables | 2011<br>pre-Modulator | 2015<br>IVA | 2019<br>Mixed<br>Cohort<sup>#</sup> | 2021<br>ELE/TEZ/<br>IVA |
| --- | --- | --- | --- | --- |
| Eligible patients on therapy (%) | - | 100 | 100 | 96 |
| Lung Function, ppFEV1 (%) | 63.4 | 68.3 | 70.5 | 78.8 |
| Nutrition, BMI ($kg/m^2$) | 22.3 | 23 | 23.6 | 24.8 |
| BMI < 18.5 | - | 10.1 | 10.5 | 4.9 |
| BMI >= 30.0 | - | 5.8 | 9.2 | 13.4 |
| Pulmonary Exacerbations (PEx) |  |  |  |  |
| Patients with annual PEx rates > 1 (%) | 39.7 | 41.7 | 41 | 19.5 |
| Number of Inpatient nights of Hospital Stays for PEx | 9.8 | 8.7 | 11.3 | 13 |
| Number of Inpatient Nights of Hospital IV Antibiotics for PEx | 14 | 9.3 | 14 | 13.3 |
| Sputum Microbiology Culture Positive Rates (%) |  |  |  |  |
| Mycobacteria | 80 | 79.7 | 87.7 | 70.7 |
| *Pseudomonas aeruginosa* | 79.1 | 65.3 | 57.7 | 46.8 |
| *Staphylococcus aureus* | 73 | 82 | 70 | 53.2 |
| Methicillin-resistant *Staphylococcus aureus* | 43.3 | 41.7 | 35.9 | 27.3 |
| *Haemophilus influenzae* | 26 | 18 | 26 | 27.3 |
| *Stenotrophomonas maltophilia* | 11.9 | 12.5 | 16.7 | 3.9 |
| *Burkholderia cepacia* Complex | 9 | 8.3 | 7.7 | 0 |
| *Achromobacter* | 1.5 | 1.4 | 11.5 | 1.3 |
| Nontuberculous Mycobacteria | 7.7 | 3.6 | 17.5 | 3.4 |
| Fungal *Aspergillus* | 6 | 12.5 | 11.5 | 5.2 |

<sup>#</sup>2019 mixed cohort of IVA, LUM/IVA, TEZ/IVA, and ELE/TEZ/IVA is the year when 100% of the CF individuals at the Adult CF Program that were eligible for each therapy were prescribed
and taking the four modulators. PEx: pulmonary exacerbations, IV: Intra Venous, BMI: body mass index

**Table 2.**
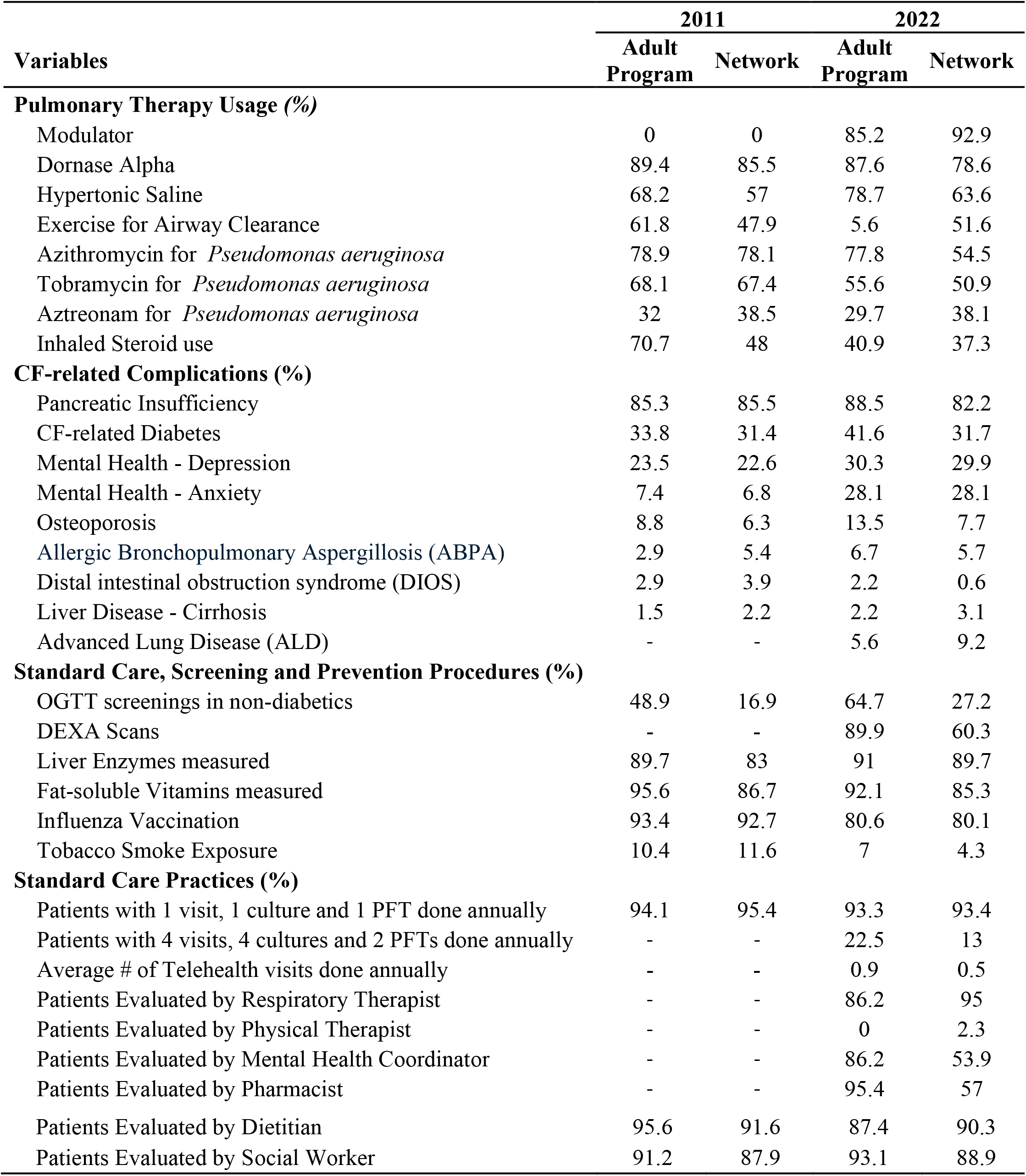
Baseline clinical characteristics and standard care practices in pre-modulator year 2011 and 2022 at Adult CF Program versus network level.

### Microbiology Trends

Over the decade, notable reductions in chronic airway infections were observed (Figure 2). The prevalence of P.a. decreased from 79.1% in 2011 to 46.8% in 2021, and Staphylococcus aureus declined from 73% to 53.2%. MRSA positivity decreased from 43.3% to 27.3%, while Mycobacterium species dropped from 80% to 70.7%. These findings suggest improved airway microbiology and reduced pathogen burden associated with CFTR modulator use, particularly during the ELE/TEZ/IVA era. Table 2 demonstrates that ELE/TEZ/IVA was associated with the greatest reductions in overall infection rates compared with IVA or mixed modulator cohorts.

**Figure 2.**
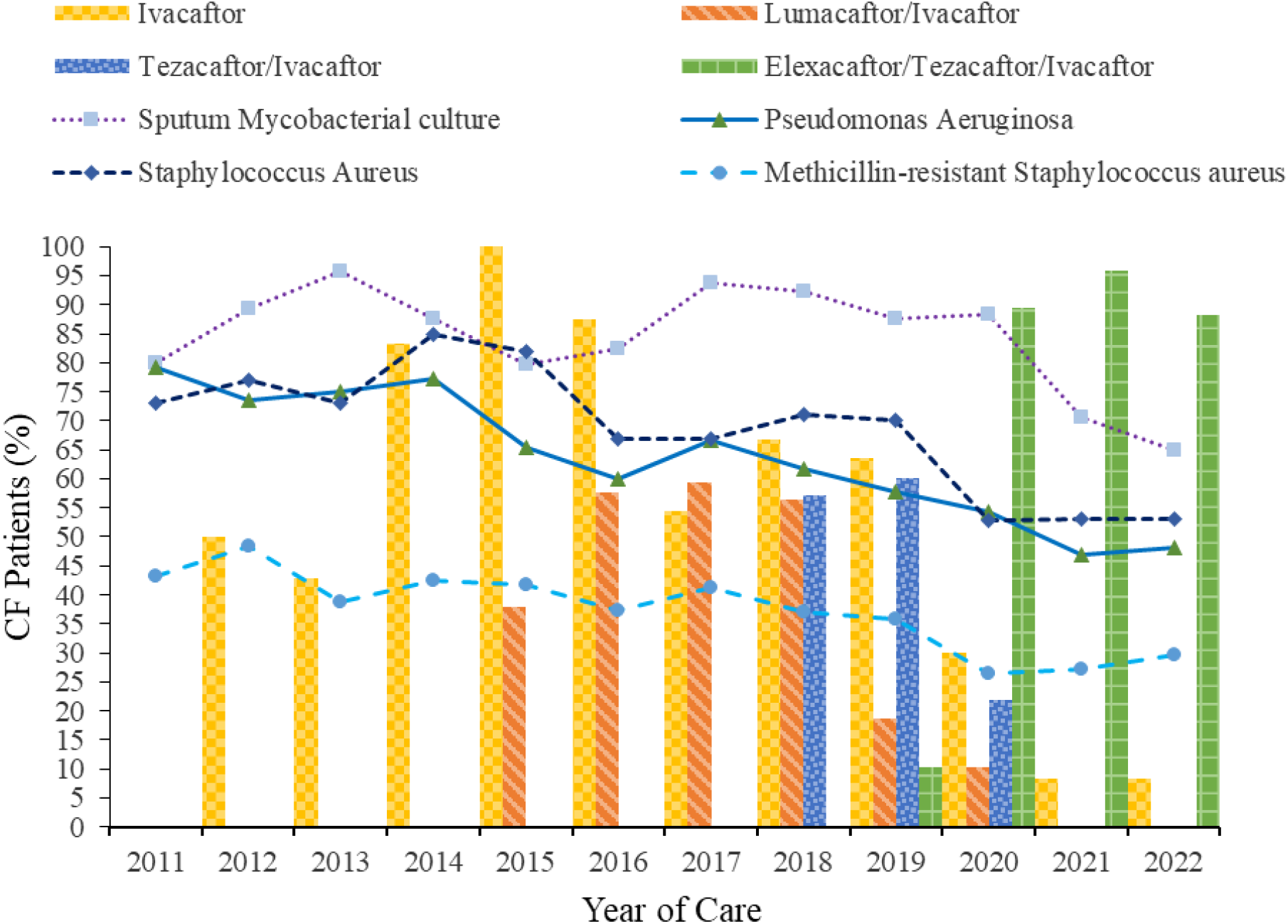
Comparative effectiveness of CFTR modulators on microbiology outcomes in adults at our Adult CF Program, 2011–2022. Vertical bars indicate the years of ivacaftor (IVA), lumacaftor/ivacaftor (LUM/IVA), tezacaftor/ivacaftor (TEZ/IVA), and elexacaftor/tezacaftor/ivacaftor (ELE/TEZ/IVA) availability. Longitudinal trends in culture positivity rates for prominent pathogens are shown as lines with markers.

### Pulmonary Exacerbations

The proportion of patients experiencing more than one pulmonary exacerbation (PEx) annually declined significantly, from 39.7% in 2011 to 19.5% in 2021 (Figure 3). Table 2 shows rates rose slightly to 41.7% in 2015, remained stable at 41% in 2019, and declined substantially following widespread ELE/TEZ/IVA use, highlighting the modulator’s effectiveness in reducing exacerbation frequency.

**Figure 3.**
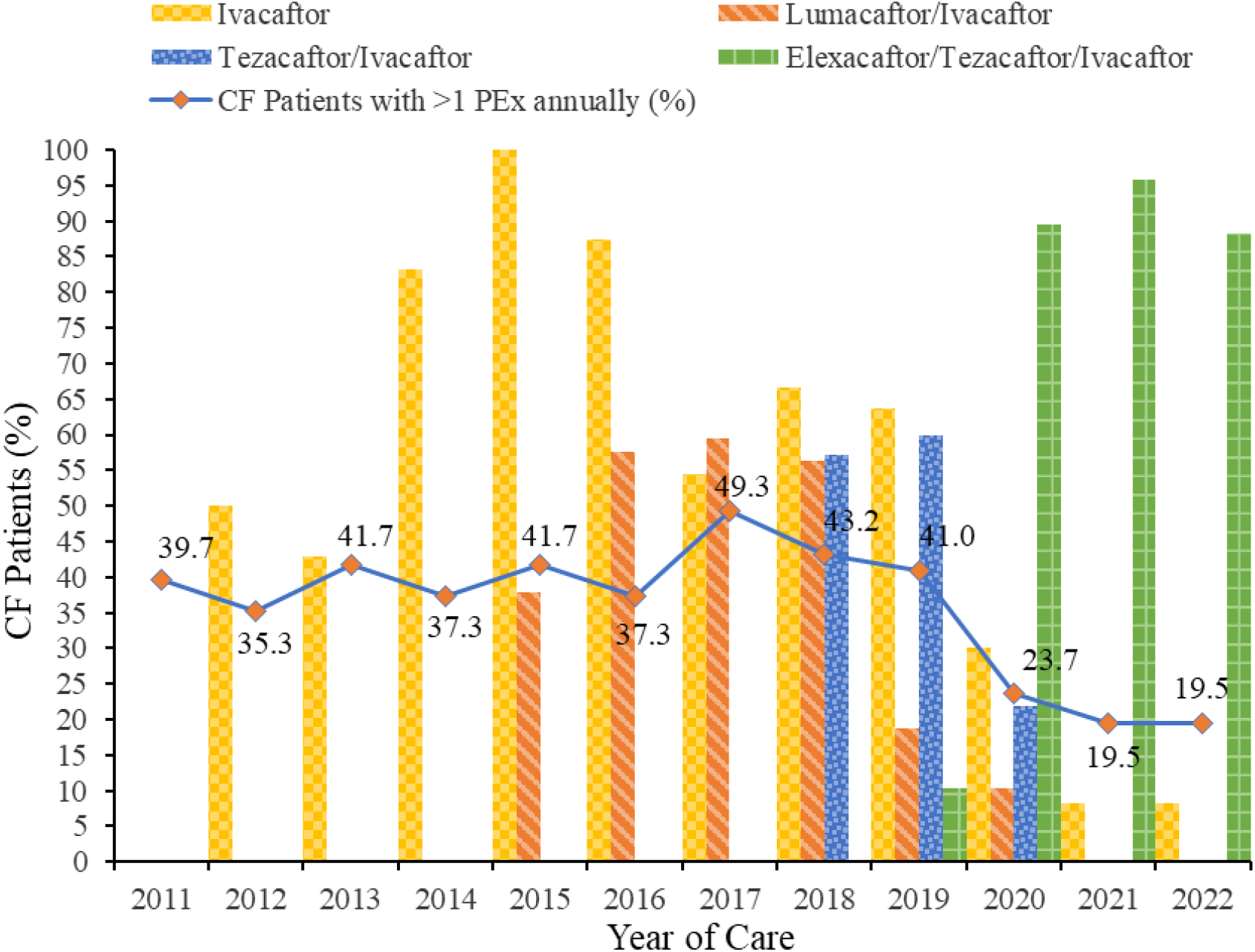
Comparative effectiveness of CFTR modulators on pulmonary exacerbation outcomes in adults at our Program, 2011–2022. Vertical bars indicate the years of ivacaftor (IVA), lumacaftor/ivacaftor (LUM/IVA), tezacaftor/ivacaftor (TEZ/IVA), and elexacaftor/tezacaftor/ivacaftor (ELE/TEZ/IVA) availability. Longitudinal trends in the percentage of adults experiencing more than one pulmonary exacerbation annually are shown as a line with markers. PEx: pulmonary exacerbations

### Network Level Performance Variations

The CFFPR Annual Program-Specific Reports provide program-level performance metrics compared with all other adult CF programs in the network, enabling benchmarking and identification of opportunities for quality improvement over the past decade. Figure 4 illustrates trends in key clinical outcomes and care processes from 2011 to 2022. Median ppFEV_1_ in our adult CF cohort was approximately 63% in the pre-modulator baseline year 2011, similar to the network median, and increased to about 81% by 2022 at both the program and network levels (Figure 4A). Median BMI values were comparable at baseline (approximately 22.0) and increased to about 24.0 in 2022 (Figure 4B).

**Figure 4.**
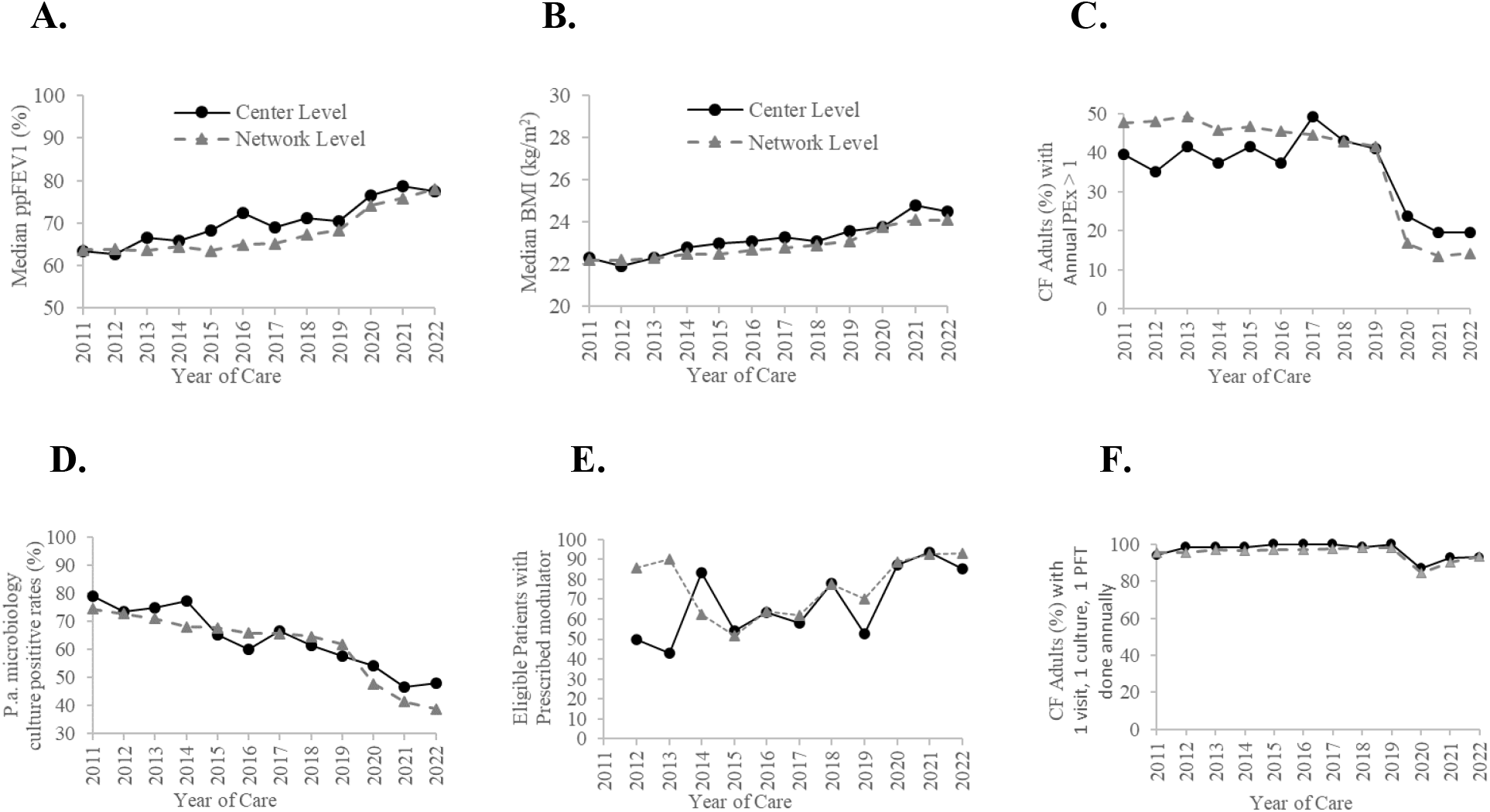
Adult CF Program versus network benchmarking using Cystic Fibrosis Foundation Patient Registry Program-Specific Annual Reports (2011–2022). Trends illustrate variations in clinical outcomes, modulator use, and adherence to guideline-based care. Program-level performance is shown with solid marker lines; network-level performance is shown with dotted marker lines. Panels display: **A**. Median ppFEV_1_ **B**. Median BMI **C**. Percentage of adults experiencing more than one PEx annually **D**. Percentage of adults with *P*.*a*. positive cultures **E**. Percentage of eligible adults receiving modulator therapy, and **F**. Percentage of adults with at least one annual visit, one respiratory culture and one PFT done as standard care. PEx: pulmonary exacerbations, ppFEV_1_: percent predicted forced expiratory volume in 1 second, BMI: Body Mass Index, P.a.: *Pseudomonas aeruginosa*

The proportion of patients experiencing more than one pulmonary exacerbation (PEx) annually decreased from 39.7% at our program in 2011 (47.7% at the network level) to 19.1% at the program and 14.2% across the network in 2022 (Figure 4C). Rates of Pseudomonas aeruginosa–positive respiratory cultures decreased from 79.1% at our program in 2011 to 49.4% in 2022, and from 74.5% at the network-level in 2011 to 38.8% in 2022 (Figure 4D). By 2022, 92.9% of eligible adults at the network level and 85.2% at our program were prescribed modulator therapy (Figure 4E).

Finally, the proportion of CF patients completing at least one annual routine clinic visit, one respiratory culture, and one spirometry test remained high across our program and all network clinics, ranging from approximately 95% in 2011 to 93% in 2022 (Figure 4F).

## DISCUSSION

In this study we utilized CFF Patient Registry data to examine decade-long trends in CFTR modulator therapy usage and their effects on clinical outcomes, as well as adherence to care guidelines, at both Adult CF Program and the broader CF Care Center Network. These results demonstrate that modulator therapies are associated with improved CF-related clinical outcomes compared to the pre-modulator year in 2011. IVA showed modest effectiveness, while the next-generation therapy ELE/TEZ/IVA was associated with the most substantial improvements in lung function (ppFEV1), BMI, microbiology profiles, and pulmonary exacerbation rates among adult CF patients. Program-level performance variation analysis over the past decade indicated that our program’s outcomes and adherence to CFF care guidelines were closely aligned with network-level performance, reflecting high-quality, standardized care.

An important limitation of this study is that CER using CFFPR data are subject to limitations such as missing or incomplete data, potential confounding, and secular trends in care delivery, which must be considered in interpretation. Despite these challenges, CER remains a powerful approach to inform clinical decision-making and guide quality improvement efforts.

While CF has been extensively studied in North America and Europe through national registries, the situation is markedly different in low- and middle-income countries. Despite an estimated burden of tens of thousands of cases, fewer have been formally diagnosed or reported, reflecting gaps in awareness, diagnostic infrastructure, and access to specialized care. In such settings, registry development and benchmarking become particularly important, as they not only capture the true disease burden but also inform standards of care where resources are limited. Furthermore, registries serve as critical tools for advocacy by providing policymakers with real-world data on disease prevalence and outcomes.The findings of our registry-based analysis underscore the broader role of patient registries beyond the United States that can transform care delivery with systemic data collection. The US based well-established registeries highlight the need for international collaboration and knowledge sharing to achieve equitable improvements in survival and quality of life for people living with CF worldwide.

Access to large, real-world datasets enables observational and CER and supports quality improvement initiatives by efficiently using existing clinical data. Registry-based studies overcome limitations inherent to clinical trials, such as high costs, restricted patient populations, and limited generalizability. By providing timely, longitudinal data, patient registries facilitate collaboration among patients, families, and clinicians, ultimately contributing to improved health outcomes, informed clinical decision-making, and advancement of CF care standards.

## CONCLUSIONS

Patient-centered care in CF has been significantly advanced through the use of patient registries. The CFF Patient Registry provides accessible, real-world data that benefit patients and families by improving understanding of disease progression and treatment effects at an individualized level. CF care centers gain insights into their patient populations, clinical outcomes, and adherence to care guidelines. Benchmarking program-level performance against the broader CFF Care Center Network enables data-driven decision-making and supports targeted quality improvement initiatives.

Registry-based CER using CFFPR data offers a robust resource to evaluate the impact of different therapies on CF-related clinical outcomes. Longitudinal registry data not only inform best practices within individual programs but also establish national and international benchmarks for care. By fostering collaboration and knowledge-sharing across programs and countries, patient registries amplify the potential to develop innovative strategies for optimizing care in rare, progressive diseases such as CF.

## Data Availability

All data produced in the present work are contained in the manuscript and the references for CF Patient Registry Program Specific Reports.

## Source(s) of Support

This study did not receive any funding.

### Ethical Approval

Ethical approval for this study was obtained from the Institutional Review Board under the ‘Research-Not Human Subject Research’ regulation.

## Acknowledgement

We would like to thank our Cystic Fibrosis patients and their families for consenting to register in the Cystic Fibrosis Foundation Patient Registry.

